# Baseline Clinical, Hormonal and Molecular Markers Associated with Clinical Response to IL-23 Antagonism in Hidradenitis Suppurativa: A Prospective Cohort Study

**DOI:** 10.1101/2023.02.22.23286201

**Authors:** A Flora, EK Kozera, R Jepsen, K Gill, J Xu, JW Frew

## Abstract

**Background:** Hidradenitis Suppurativa is a complex inflammatory disease in which predicting therapeutic response remains challenging. IL-23 interacts with sex hormones but the relationships between the two in HS remains uninvestigated.

**Objectives:** To assess whether baseline clinical, hormonal, or molecular markers are associated with clinical response to IL-23 antagonism with Risankizumab in Hidradenitis Suppurativa.

**Methods:** 26 individuals with Hurley Stage 2/3 disease were administered Risankizumab 150mg Week 0,4,12. Baseline sex hormones and skin biopsies were taken. Clinical response at Week 16 assessed by the HiSCR, and differences between responders and non-responders assessed.

**Results:** 18 of 26 participants achieved HiSCR50 at week 16 (69.2%). Clinical response to IL-23 antagonism was associated with male gender, elevated total serum testosterone, and decreased levels of FSH. Stratification by clinical responders/non responders identified differentially expressed genes including *PLPP4* and *MAPK10*. Immunohistochemistry identified elevated numbers of CD11c, IL-17A and IL-17F positive cells compared to non-responders. CD11c+ cells significantly correlated with serum levels of total testosterone and inversely correlated with serum FSH.

**Conclusions:** Clinical response to IL-23 antagonism in HS is associated with serum sex hormones, Th17 polarized inflammation in lesional tissue and CD11c+ cells. These potential therapeutic biomarkers require further validation in larger cohorts but may suggest potential targeted HS therapy.

## Main Text

Hidradenitis Suppurativa (HS) is a complex, heterogeneous inflammatory disease in need of novel therapies^1^. Currently, Adalimumab is the sole licensed biologic therapy for HS, and only achieves clinical response (as measured by the Hidradenitis Suppurativa Clinical Response outcome measure) in 60% of patients^2^. Mechanistically, HS is associated with significant polarization of the Th17 immune axis with significant dysregulation of cytokines including IL-17A^3^, IL-17C^4^, IL-17F^3,5^ and IL-23^3^. Agents targeting the Th17 immune axis have been explored in HS^6^ including clinical trials^7^, but further development has been halted due to lack of positive results^8^. Two separate IL-23 antagonists (Risankizumab and Guselkumab) have been withdrawn from further development after a lack of positive results in Phase 2 studies^8^, with no significant difference in clinical response when compared to placebo^8^(Supplementary Table 4). Isolated case reports and case series however, report benefit in some patients^6,9^

HS also has a significant hormonal component to disease pathogenesis^10^. The link between sex hormones and the Th17 immunological axis is incompletely understood^11^. Perimenstrual disease flares, remission during pregnancy and the effect of comorbidities such as obesity and insulin resistance on hormone levels suggest sex hormones play an important role in modulating inflammation in HS^12^. Hormonal therapies such as Spironolactone, oral contraceptives and 5-α reductase inhibitors such as finasteride and dutasteride are used, primarily in female patients, with variable levels of efficacy^12,13^.

The Th17 immune axis is known to interact with sex hormones through IL-23^14,15^. IL-23 interacts with non-canonical androgen receptor signaling pathways, modulating inflammation in epithelial tissues and monocytes/macrophages^14,15^. Monocyte activation and development into macrophages are altered in the presence of tissue estrogens, (specifically E2 Estradiol) both in in vivo and in vitro investigations^14,15^. Monocytes and macrophages are known to be central players in the pathogenesis of HS^16-19^, produce IL-23, and are proposed to interact with T cells, B cells and Neutrophils to direct and orchestrate chronic inflammation in HS^16-19^.

Recent Phase 2 studies of IL-23 antagonism in HS have suggested disparate clinical responses based upon participant gender, with primarily male cohorts demonstrating higher rates of clinical response to IL-23 antagonism compared to primarily female cohorts^8^. This would be consistent with sex hormones influencing the role of IL-23 directed inflammation in HS, however clinical and mechanistic evidence to support this hypothesis is currently lacking.

The aim of this prospective cohort study was to assess whether baseline clinical, hormonal or molecular markers are associated with clinical response to IL-23 antagonism with Risankizumab in Hidradenitis Suppurativa.

## METHODS

26 individuals with dermatologist diagnosed HS (based upon the modified Dessau criteria^20^) were included in this cohort study. All individuals had Hurley stage 2 or 3 disease and a minimum of 5 inflammatory lesions (abscesses and nodules). Inclusion and Exclusion criteria are presented in supplementary table 1. Clinical data including demographics, smoking status, body mass index (BMI), family history of HS, and diagnosed insulin resistance were collated (Supplementary Table 2). Disease severity was assessed using Hurley staging and lesion counts using the international hidradenitis suppurativa severity score (IHS4) outcome measure^2^. Baseline blood work including serum total and free testosterone, follicular stimulating hormone (FSH), luteinizing hormone (LH), sex hormone binding globulin (SHBG) as well as C reactive protein (CRP) levels. All participants were administered Risankizumab at psoriasis dosing of 150mg week 0,4 and 12. The primary outcomes of interest was clinical response as measured by the Hidradenitis Suppurativa Clinical Response (HiSCR^2^) at week 16. Additional outcomes included the IHS4, as well as HiSCR75 (defined as a 75% reduction in Abscess and Nodule count without an increase in abscesses or draining tunnels), and HiSCR90 (defined as a 90% reduction in Abscess and Nodule count without an increase in abscesses or draining tunnels) as used in the recent Bimekizumab Phase 2 clinical trial in HS^21^. Clinical Responders were defined as individuals who achieved HiSCR at Week 16. Non-Responders were defined as those who did not achieve HiSCR 50 at Week 16.

### Skin Biopsy Collection and Processing

Lesional, peri-lesional and non-lesional skin biopsies were taken prior to the commencement of Risankizumab using previously described standardized lesion and site methodologies^22^. Each 6mm biopsy specimen was bisected, with one section immediately placed in RNA later and frozen at −80 degrees Celsius until processing for RNA extraction. The other section was placed in OCT medium and frozen at −80 degrees Celsius and processed for Immunohistochemistry.

RNA was extracted using the Qiagen RNEasy kit then eluted in 50ug of RNA-ase free water. RNA was processed and analysed using the Nanostring nCounter Fibrosis 2.0 multiplex gene expression assay (gene list in Supplementary Table 3). nCounter technology uses a pair of gene-specific probes – a capture probe and a reporter probe – with each aligning to their target RNA by complementary base pairing. The reporter probe contains a target-specific fluorophore, and readout is via an imaging platform that identifies and quantifies probe complexes^23^.

Tissue sections in OCT were cut into 5.0μm sections and stained for the following proteins of interest: IL-17A (Thermofisher 14717982, 1:100), IL-17F (Thermofisher PA5115403 1:100), IL-23p19 (Thermofisher PA520239 1:100) and Cd11c (Proteintech KHC0017) Appropriate secondary antibodies (Abcam goat anti-mouse Ab6789 1:2000), Abcam goat anti rabbit Ab6721 1:200) were used with DAB chromophore. IHC quantification was undertaken using semiquantitative measurement by 2 experienced independent raters (AF, JWF), with any disagreement mediated by a third author.

### Statistical Analysis

Power calculation was undertaken to determine the significance of findings. Assuming a clinical response rate of 40% similar to clinical trial data^8^, a sample size of 26 participants will allow detection of significant change in two markers (with log fold change greater than 1.5) with power greater than 80% if one tailed significance T-tests is performed.

Descriptive statistics were used to collate all demographic and disease severity data in the included patients. Differences between groups (Responders/Non responders) were analyzed using Chi Squared analysis for binary/dichotomous variables and the Wilcoxon Rank sum test for continuous variables. P<0.05 was considered statistically significant and adjustment for multiple comparisons was made using the Benjamini Hochberg procedure. All statistical analysis was completed in Graphpad Prism (9.4.1)

### Analysis of Serum Sex Hormones

Serum sex hormones were measured by the same laboratory across all individuals. All pre-menopausal female participants underwent baseline blood tests during the first 3 days of their menstrual cycle to account for cyclical variation. As per the exclusion criteria-no women in this cohort study were on active hormonal contraception. Differences between groups were analyzed using Wilcoxon Rank sum test for continuous variables. P<0.05 was considered statistically significant and adjustment for multiple comparisons was made using the Benjamini-Hochberg procedure.

### Immunohistochemistry quantification

IHC staining underwent semiquantitative analysis using previously published methods^24^. Differences between groups were analyzed using the Wilcoxon rank sum test with adjustment for multiple comparisons was made using the Benjamini Hochberg procedure.

### Nanostring NCounter Analysis

Extracted RNA was analysed using the NCounter system (Nanostring) using the Human Fibrosis V2.0 gene panel. Raw data was processed using the Nanostring Nsolver (version 4.0.70) analysis software using quality control and normalization procedures derived from the NormqPCR R package as previously described^25^. Differentially expressed genes (DEGs) were defined as >1.5 Log2Fold change with a false discovery rate<0.05 and p value<0.05.

## RESULTS

### Clinical response to IL-23 antagonism was significantly associated with male gender, insulin resistance and serum sex hormones

Eighteen of the twenty-six included participants were classified as responders (69.2%), having achieved HISCR at Week 16. Responders demonstrated dramatic reduction in the number of clinically apparent nodules and abscesses, but also draining tunnels as well as diffuse erythema, pain and swelling (Supplementary Figure 1). The demographic and disease characteristics of the included patients (including Responders and Non-Responders) is presented in Table 1. A statistically significant difference in the proportion of male participants in the responder category when compared to the non-responder category as measured by the Chi Squared test. (77.8% vs 12.5% p<0.01). There was no significant difference between responders and non-responders with regards to age, BMI, or Hurley stage (Table 1). Significant differences in total testosterone, FSH and LH were also identified between responders and non-responders. (Table 1, Figure 2).

**Table 1:** Table of summary demographic and disease data of participants in this cohort study.

| Variable | Total (n=26) | (HiSCR50) |  | P-Value |
| --- | --- | --- | --- | --- |
|  |  | Responders<br>(18/26, 69.2%) | Non-Responders<br>(8/26, 30.8%) |  |
| Gender n (%) | Male: 15 (57.7%)<br>Female: 11 (42.3%) | Male 14/18 (77.8%)<br>Female 4/18 (22.2%) | Male: 1/8 (12.5%)<br>Female: 7/8 (87.5%) | P<0.01 |
| Age<br>(Median) | Median=31.5 | Median= 31 | Median=40 | P=0.08 |
| BMI<br>(Median) | Median=28 | Median=28 | Median= 36.5 | P=0.11 |
| Insulin<br>Resistance<br>n, (%) | 4/26 (15.4%) | 4/18 (22.2%) | 0/8 (0%) | P<0.001 |
| Hurley Stage<br>(n,%) | Hurley 2:<br>13/26 (50%)<br>Hurley 3:<br>13/26 (50%) | Hurley 2:<br>7/18 (38.9%)<br>Hurley 3:<br>11/18 (61.1%) | Hurley 2:<br>6/8 (75%)<br>Hurley 3:<br>2/8 (25%) | P=0.06 |
| Baseline IHS4<br>Score<br>(Median) | 42 | 40 | 41 | P=0.87 |
| Median<br>Serum Total<br>Testosterone | 9.7nmol/L | 20.3 nmol/L | 1.0 nmol/L | P<0.001 |
| Serum FSH | 6.2 mIU/mL | 3.2 mIU/mL | 10 mIU/mL | P<0.001 |
| Serum LH | 5.45 IU/L | 4.5 IU/L | 16.5 IU/L | P=0.01 |
| Serum CRP | 12.3 mg/L | 13.0 mg/L | 11.2 mg/L | P=0.36 |

**Figure 2:**
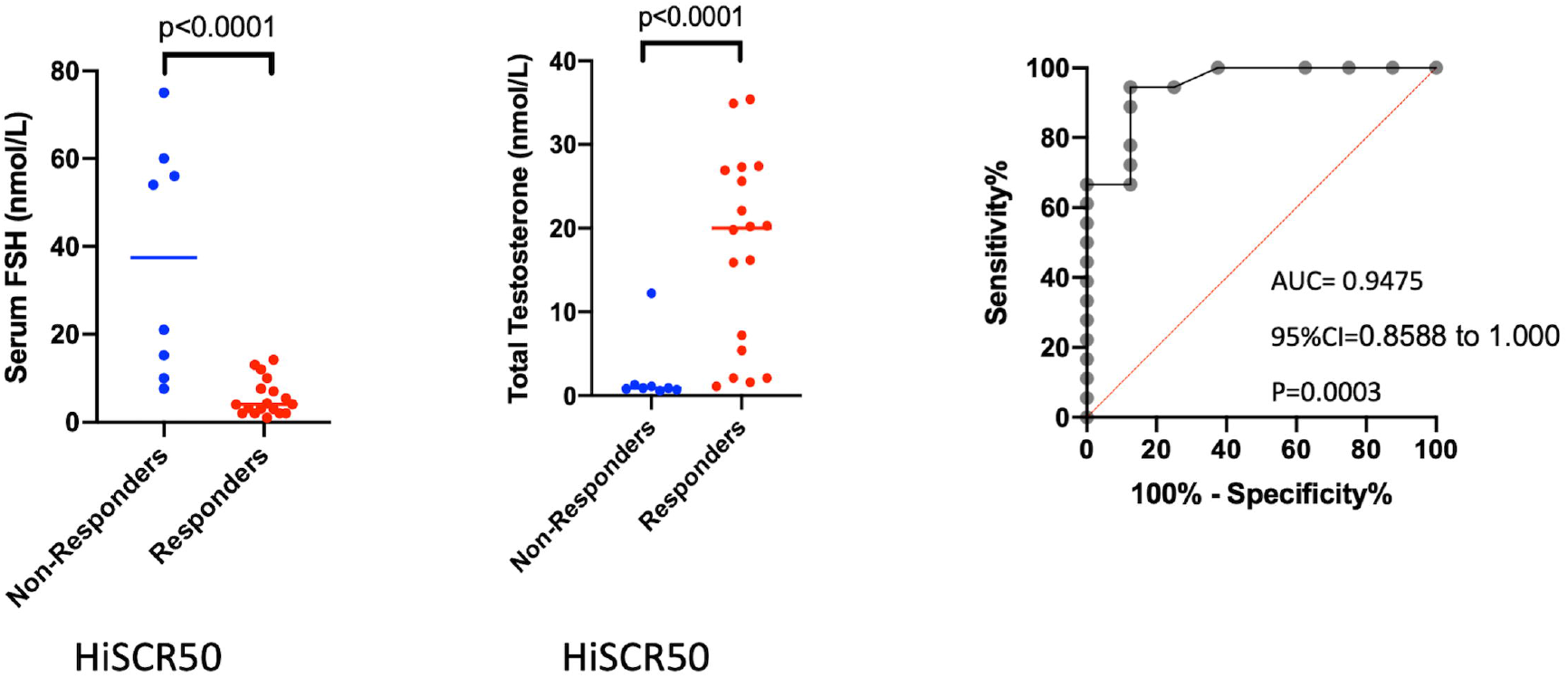
Comparison of Baseline Serum Testosterone and Serum FSH levels in Responders and Non-Responders (at week 16) to Risankizumab therapy for Hidradenitis Suppurativa as well as receiver operating curve for testosterone and HiSCR50.

The univariate association of clinical response as measured by HiSCR (hereafter termed HiSCR50) was maintained when deeper measures of clinical response (HiSCR75 and HiSCR90) were analyzed (Supp Figures). Statistically significant differences between serum FSH and serum total testosterone were identified between HiSCR75 Responders/Non-Responders. Only serum total testosterone was statistically significant between HiSCR90 Responders/Non-Responders/(Supplementary Figures 2,3,4,5).

No statistically significant differences between HiSCR Responders/Non-Responders were seen between other hormonal measurements including sex hormone binding globulin (SHBG), free testosterone and serum luteinizing hormone (LH). (Supplementary Figures 2,3,4,5) Additionally, exploratory correlation between age and BMI did not reveal any statistically significant difference between clinical Responders and Non-Responders (Supplementary Figures 2,3,4,5). Examination of deeper levels of response including HiSCR75 and HiSCR90 also did not identify any significant differences between responders and non-responders. (Supplementary Figures 2,3,4,5)

Analysis of the relationship between clinical response and FSH/total testosterone by multiple regression analysis did not reveal any loss of association in the presence of other identified variables (Supplementary Table 6). This indicates that serum total testosterone and serum FSH are consistently associated with clinical response, as measured by HiSCR50/75/90 outcome measures, even in the presence of other demographic and hormonal variables.

### HS Lesional, Perilesional and Non-Lesional Tissue Demonstrates Dysregulation of Multiple Inflammatory and Immunological Pathways by Multiplex Gene Expression, similar to those observed in whole tissue RNA sequencing

Gene expression using multiplex gene expression (Nanostring nCounter Fibrosis 2.0 gene expression panel) identified significant dysregulation of fibro-inflammatory genes including *IL1B, CXCL8, SYK, CXCR1*) across lesional, perilesional and non lesional tissue (Figure 3). The highest differentially expressed genes (Supplementary Figure 6, 7) included genes and pathways previously identified in RNA sequencing including interferon responsive elements (*IRF1*), neutrophil activation (*CEACAM3*), B cell activity (*BLK*) and Th17 pathways (*IL17A*). Additionally, a number of novel genes and pathways were highlighted including CD8 cell exhaustion (*LAG3*), mast cell activity (*C4BPA*) and sex-hormone regulated monocyte associated genes (*PRAP1*).

**Figure 3:**
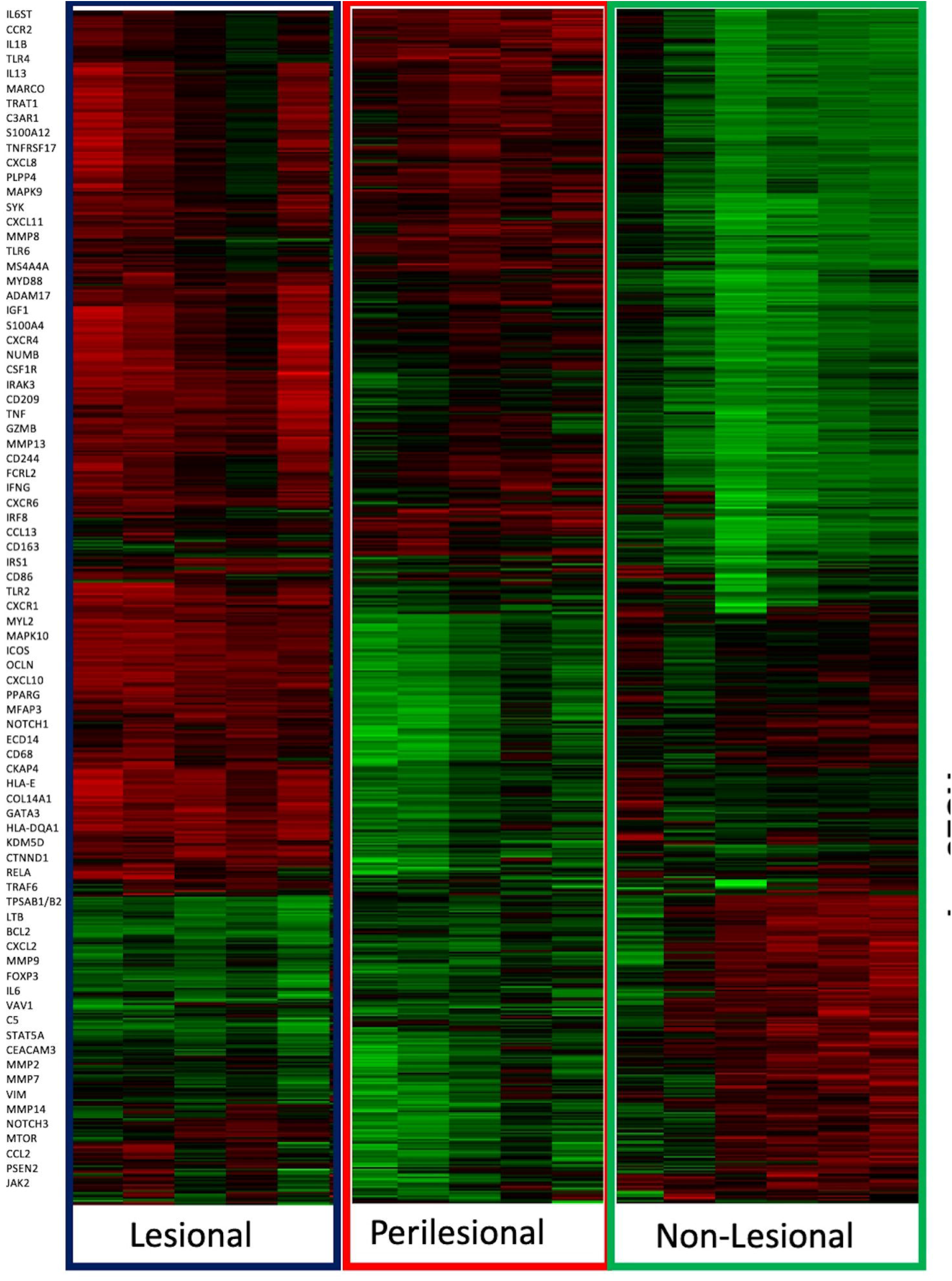
Baseline Multiplex gene expression (Nanostring) data for baseline tissue biopsies in 26 participants, stratified by lesional, perilesional and non lesional tissue.

Principal component analysis (Supplementary Figure 6) identified specific genes significantly associated with lesional tissue (including *BLK, CD44, IL6, STAT3*), peri-lesional tissue (*IL10, C5, IRF1*) and non-lesional tissue (*ILK, COLA12, IL17A*). These genes and associated cell types have been observed in whole tissue RNAseq^26,27,28^ validating the novel use of this technology in HS tissue.

### Molecular response stratified by HiSCR Responders/Non-Responders identify upregulated T cell, Th17 associated genes and pathways in clinical responders to IL-23 antagonism

Stratification of baseline tissue gene expression by HiSCR50 responders and non-responders identified a number of genes associated with clinical response to Risankizumab therapy (Figure 4).The most differentially expressed genes pertained to extra cellular matrix synthesis (*MFAP3, CYP8B1*) as well as T cells (*TRAT1*), PPAR signaling (*CYP8B1*), mast cells (*TPSAB1/B2*) and cytokine signaling (*CXCR1, LTA, FADD, OAS1*). Statistically significant dysregulated genes are listed in Figure 4). Sex-hormone modulated genes including *PLPP4* and *MAPK10* were upregulated in responders compared to non-responders. Principal component analysis identified significant clustering between responders and non-responders when stratified by HiSCR50 (Figure 4). Pathway analysis by reactome identified interleukin-17 signaling, chemokine receptors, and signaling by interleukins, Innate immune system and immune system as significant pathways in responders versus non-responders (Supplementary Table 5).

**Figure 4:**
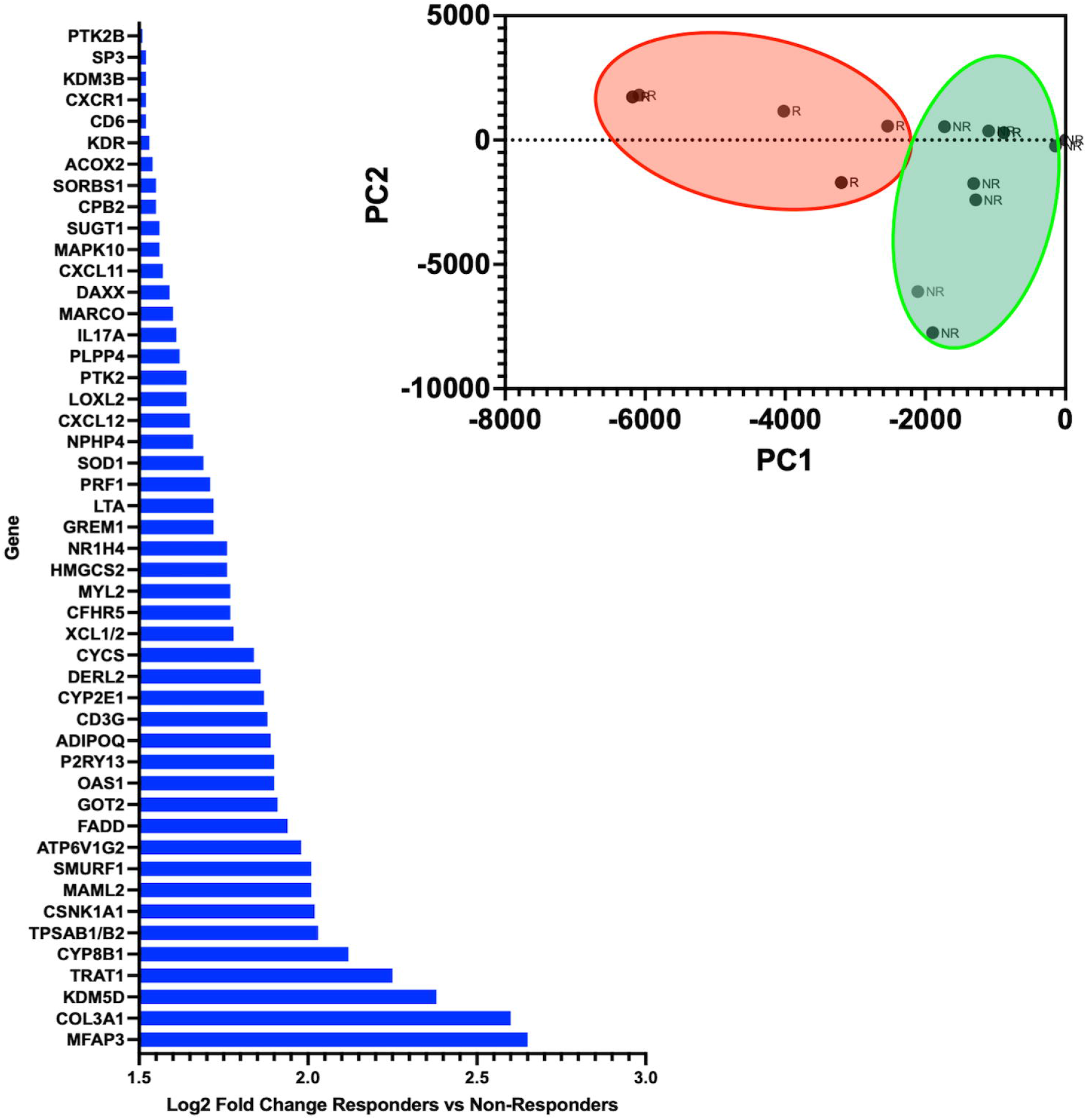
Log fold change in gene expression of select genes in responders versus non-responders in Hidradenitis Suppurativa. Statistically significant elevation in gene expression was seen in T cell, macrophage monocyte and dendritic cell genes compared to non-responders. This included IL-17A, CXCR1, TRAT1 and LTA. Principal component analysis demonstrating discrete clustering of Responders and Non-Responders based on gene expression data.

### Immunohistochemistry identifies Th17 associated proteins such as IL-17A, IL-17F and IL-23p19 as well as and CD11c+ leucocytes as associated with clinical response in HS lesional tissue

Confirmatory IHC indicates significantly increased number of CD11c+ cells and IL23p19+ cells in dermal infiltrates in lesional tissue of clinical responders (as measured by HiSCR) when compared to clinical non-responders. (Supplementary Figure 8). Semiquantitative cell counts identified elevated levels of IL-17A and IL-17F positive cells in dermal infiltrates of lesional tissue in clinical responders (Figure 5). Univariate correlation identified a significant association between Serum FSH and Serum testosterone with semiquantitative IHC cell counts of CD11c and IL23p19 positive cells (Supplementary Figure 8).

**Figure 5:**
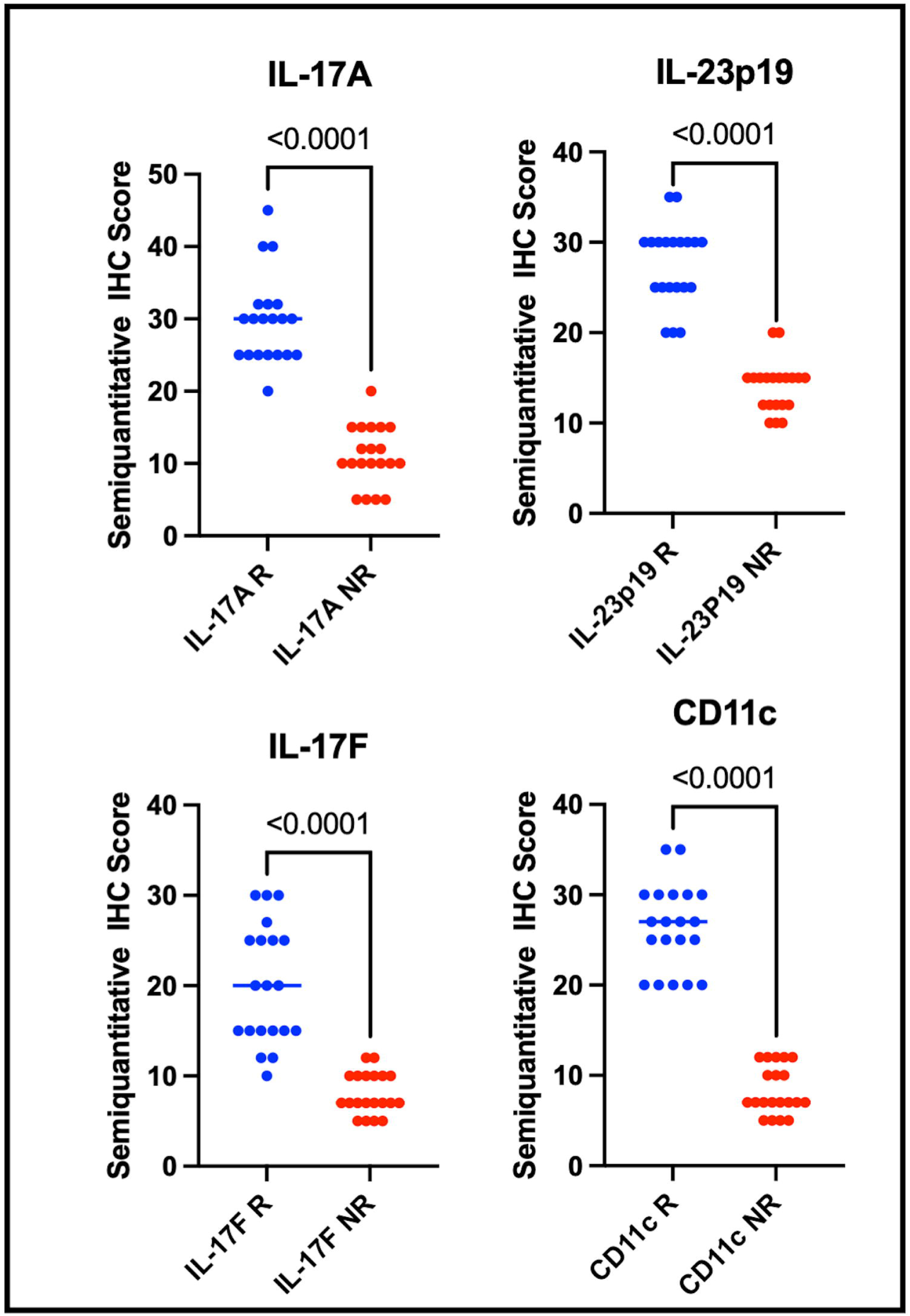
Immunohistochemical staining for IL-23p19, CD11c, IL-17A and IL-17F are associated with clinical response to Risankizumab therapy in Hidradenitis Suppurativa.

## Discussion

Clinical response to IL-23 antagonism with Risankizumab in HS is associated with baseline elevation in total testosterone and baseline suppression of FSH, as well as upregulation of sex-hormone responsive genes including *PLPP4* and *MAPK10* in lesional tissue. The greater proportion of males in this cohort (compared to other placebo-controlled trials in HS) may explain the slightly higher response rate in this cohort. Different patient factors are associated with clinical response in females as compared to males. In females, insulin resistance is also associated with clinical response suggesting a complex interplay between inflammation, sex hormones and metabolic comorbidities such as polycystic ovarian syndrome (PCOS). Additionally, responders were associated with increased numbers of baseline lesional IL23p19+ and CD11c+ cells in dermal inflammatory infiltrates as well as upregulation of Th17 inflammatory pathways. Data from this observational study suggests that sex hormones may be associated with IL-23p19 expressing CD11c+ cells in HS lesional tissue, which are associated with elevations in the Th17 immune axis and therefore clinical response to IL-23 antagonism.

This study was not designed to look at causation, and hence only associative conclusions can be drawn. It may be that serum testosterone and serum FSH are surrogate markers for other causative mechanisms which are currently unknown. What this study does provide is observational evidence that clinical response to IL-23 antagonism associates with alterations in serum sex hormones. Additionally, these hormone alterations are associated with Th17 immune axis skewing, upregulation of *MAPK10* gene expression in lesional tissue and increases in CD11c+ cell populations in lesional tissue.

Levels of sex hormones are influenced by age, the menstrual cycle, pregnancy and medication such as the contraceptive pill^29,30^. Whilst the influence of sex hormones on the activity of HS is well documented^10^, the link between sex hormones and inflammatory cytokines and chemokines in HS is not well understood. Monocyte populations are known to be altered in the setting of HS^19,26,27^ with trends toward non-classical populations observed^19,26.27^ but little data has observed differences in monocyte populations between men and women with HS.

PCOS is known to alter the populations and activity of CD11c+ monocytes^31^, and sex hormones can alter monocyte differentiation with testosterone driving a non-classical monocyte differentiation and estrogens forming a classical differentiation program^32-25^. This is manifest by alterations in CD14 and CD16 expression, however this data has not been replicated in HS. Additionally, estrogens can depress the dendritic cell stimulation of T cells, and testosterone promote inflammatory activation of monocyte-derived dendritic cells^32-35^.

Our presented results would be consistent with previous observations that sex hormones may impact the ratio of classical/intermediate/non-classical monocytes^32-25^. An overarching explanation for the observations in this study would be that elevated levels of testosterone in IL-23 clinical responders may result in preferential intermediate and non-classical monocyte polarization (via *MAPK10* and *CXCL1* gene expression^32^), manifesting in high levels of CD11c expression and Th17 immune polarization compared to IL-23 clinical non-responders.

An additional novel feature of this report is the utilization of the nCounter Nanostring gene expression system, which when used in other inflammatory dermatoses^36^ has suggested higher sensitivity than traditional RNAseq (particularly with inflammatory cytokines such as IL-17 isoforms). The similar upregulation of inflammatory targets in lesional, perilesional and non-lesional tissue when compared to other published datasets^5,27,28^ partially validates the use of this gene expression method in HS.

### Limitations

Limitations to this study include the small number of participants, the cohort nature of the study and the use of only baseline serum and tissue data as opposed to pre and post intervention sampling. The use of gene expression panel (nCounter) is reliant on a prior identified gene groups as opposed to assumption free RNAseq. However, the increased sensitivity of this technology promotes possible benefits from this over and above traditional RNA sequencing. The observations gained can only look into association not causation of sex hormones, CD11c+ populations and clinical response. All women included in this study were not on any form of contraceptive therapy. Hence the potential beneficial or detrimental role of exogenous hormones was not able to be assessed. Additionally, levels of tissue aromatase (which can convert androgens to estrogens) was not assessed. This may account for disparity between serum levels of sex hormones and their local action in tissue.

Future directions would include looking at pre and post interventional studies to potentiate response to IL-23 antagonisms (through the use of hormonal manipulation), the role of sex hormones in monocyte subsets in inflammatory skin disease, as well as exploring the effect of the OCP other hormonal agents (such as Spironolactone) in concert with IL-23 antagonism in HS.

## Conclusions

Overall, individuals demonstrating a clinical response to IL-23 antagonism in HS exhibit elevated levels of total serum testosterone and elevated CD11c+ cell numbers in lesional tissue. Elevated expression of *IL17A, PLPP4* and *MAPK10* genes were observed in clinical responders suggesting a potential link between sex hormones and modulation of monocyte differentiation in HS tissue. These potential therapeutic biomarkers require validation in larger cohorts, and the influence of hormonal medications in potentiating or facilitating clinical response to IL-23 antagonism may be an interesting future direction of translational research.

## Data Availability

All gene expression data is available under GEO Accession GSE214820

## Data Availability

All gene expression data is available under GEO Accession *GSE214820*

## Ethics Approval Statement

This study was approved by the Human Research Ethics Committee of South Western Sydney Local Health District.

## Contributor Statement

AF: Investigation, Methodology, Writing, Revision

EKK: Methodology, Writing, Revision

RJ: Investigation, Methodology, Writing, Revision

JX: Methodology, Writing, Revision

KG: Methodology, Writing, Revision

JWF: Conception, Funding, Supervision, Investigation, Methodology, Writing, Revision

## Data Availability Statement

The datasets used in this manuscript are publicly available through Gene Expression Omnibus (GEO) via accession number GSE214820

## Supplementary Figures

Supp Figure 1: Representative clinical photographs of response to Risankizumab therapy at Week 16

Supp Figure 2: Comparison of FSH and Total Testosterone between Responders and Non responders stratified by HiSCR75 and HiSCR90.

Supp Figure 3: Comparison of Sex Hormone Binding Globulin (SHBG); Luteinizing Hormone (LH), and Body Mass Index (BMI) between Responders and Non-Responders stratified by HISCR50

Supplementary Figure 4: Comparison of Age, Sex Hormone Binding Globulin (SHBG); Body Mass Index (BMI), Luteinizing Hormone (LH), and free testosterone between Responders and Non-Responders stratified by HISCR75

Supplementary Figure 5: Comparison of Sex Hormone Binding Globulin (SHBG); Age, free testosterone, Luteinizing Hormone (LH), Body Mass Index (BMI), and between Responders and Non-Responders stratified by HISCR90

Supplementary Figure 6: Principal Component Analysis demonstrating clustering of lesional and perilesional tissue samples distinct from non lesional samples with highlighted associated genes. Log2Fold change of genes in lesional, perilesional and non-lesional tissue compared to site-matched healthy controls.

Supplementary Figure 7: Heatmap with hierarchical clustering demonstrating discrete clustering of Responders and Non-Responders to Risankizumab in Hidradenitis Suppurativa.

Supplementary Figure 8: Representative immunohistochemistry for CD11c and IL23p19 in responders and non responders to Risankizumab in Hidradenitis Suppurative. Additionally, correlation between Serum FSH and Serum Testosterone with CD11c+ IHC score.

## Supplementary Tables

Supp Table 1: Inclusion, Exclusion Criteria

Supp Table 2: Table of Individual Patient Characteristics of participants

Supp Table 3: nCounter Human Fibrosis V2.0 Gene List

Supp Table 4: Data from the NOVA Trial adapted from NCT03628924

Supp Table 5: Reactome Pathway Analysis of Responders to Risankizumab Therapy

Supp Table 6: Logistic Regression analyses for HiSCR50/HiSCR75 and HiSCR90

